# The impact of Recovery College enrolment on health service use and patient outcomes: retrospective matched cohort study using routinely collected data

**DOI:** 10.64898/2025.12.09.25341905

**Authors:** Amy Ronaldson, Thomas Allen, Ioannis Bakolis, Richard Emsley, Tesnime Jebara, Yasuhiro Kotera, Danielle Dunnett, Simran Kaur Takhi, Merly McPhilbin, Jonathan Simpson, Agnieszka Kapka, Helen Killaspy, Daniel Hayes, Mariam Namasaba, Sara Meddings, Amelia Jewell, Kirsty Giles, Lisa Brophy, Dora Shergold, Jason Grant-Rowles, Peter Bates, Rachel A Elliott, Claire Henderson, Mike Slade

## Abstract

**Background:** Recovery Colleges (RCs) support recovery through adult education, with preliminary evidence of positive effects on a range of outcomes. This study examined associations between RC enrolment and mental health service use at an index mental health provider, use of other National Health Service (NHS) hospital services for all causes, associated costs, and service user outcomes.

**Methods:** Our retrospective matched cohort study used a controlled before-and-after design. We used linkage with electronic health records to identify all mental health service user students enrolled at one RC. Students were matched with non-student service user controls on sociodemographic and clinical variables using caliper matching. Impacts of RC enrolment on service use were assessed using negative binomial regression models at six-month, 12-month, and five-year post-enrolment. People with lived experience were involved in the design, conduct, and reporting of this study.

**Outcomes:** Our sample comprised 1 435 students and 4 665 controls. We observed decreases in several types of mental health service use in students relative to controls at six months (e.g. adjusted Incidence Rate Ratios [aIRRs] for inpatient admissions 0·56, 95%CI 0·30 to 0·64) and 12 months (aIRR 0·60, 95%CI 0·44 to 0·81). At 12 months, students showed a £5 028 (95%CI -£8 223 to -£1 834) greater reduction in total costs per student compared with controls. This indicates that RCs offer an 8·4:1 financial return on investment. Students also showed relative reductions in all-cause hospital bed days at six months (aIRR 0·53, 95%CI 0·35 to 0·81) and 12 months (aIRR 0·66, 95%CI 0·46 to 0·96), with a £412 (95%CI -£1 085 to -£260) greater reduction in associated total costs at 12 months. Among students, reductions in Health of the Nation Outcome Scale (HoNOS) scores indicated consistent improvement in functioning over time.

**Interpretation:** Mental health service users who enrol in a RC have reduced subsequent mental and all-cause healthcare use, and reduced service-related costs compared with matched service users not using a RC. Service user outcomes are also improved.

**Funding:** National Institute for Health and Care Research.

**Research in context:** *Evidence before this study:* Since the first one opened in England in 2009, Recovery Colleges (RCs) have spread globally. A 2025 review collating 2013-2024 evidence (64 papers) identified 11 studies investigating outcomes and four investigating service use. Quantitative evaluation of outcomes has used pre-post designs to investigate the impact of RCs on components of recovery, finding consistent evidence of benefit in relation to a number of outcomes including wellbeing, empowerment, hope, and social inclusion. Service use studies have indicated benefits from RC attendance, including increased employment and reduced hospital admissions and bed days, with preliminary evidence of associated cost savings. However, across all studies the evidence quality is low, with most outcome studies using small samples (mostly <100 students) and none using a separate control group. Consequently, change due to other factors such as measurement error or time cannot be discounted, so causation cannot be established.

*Added value of this study:* This is the largest study of its kind which utilises a methodologically rigorous approach to investigate the impact of RCs on service use, costs and outcomes. In 6 100 people, we identified a consistent positive impact for service user students, compared with optimally matched service user non-students, in relation to mental health service use at an index mental health provider (especially in-patient admissions) and wider all-cause hospital service use (especially bed days) at 12 months post-enrolment, resulting in relative cost savings for service users who are students compared with those who are not. Furthermore, we showed a relative beneficial impact for students on functioning consistently up to five years after RC enrolment.

*Implications of all the available evidence:* The evidence base for supporting RCs is significantly strengthened. Mental health service users who are students at RCs are likely to benefit, both in terms of clinical outcomes and reduced service use, compared to similar people not using the RC. Significant cost savings also arise, which we estimate as an 8·4:1 financial return on investment. Our study evidence supports ongoing investment in RCs with significant return on investment, especially in England.

## Introduction

Recovery Colleges (RCs) aim to support the personal recovery of people with mental ill health through coproduced adult education, rather than through treatment, where people have the role of student rather than patient.^1^ Personal recovery is described as the subjective process of taking control of one’s life and illness, having optimism for the future, and taking personal responsibility for one’s own recovery.^2^ This is in line with the current vision of the World Health Organization, who have recognised recovery-oriented practice as the guiding vision for mental health systems internationally.^3^

The two foundational principles of RCs are adult education and coproduction.^4^ Students, facilitators, and educators share experiences, knowledge, and skills to develop students’ strengths through participation in recovery-oriented courses.^5^ Since their inception in 2009, RCs have gained widespread momentum; a global survey identified 221 RCs in 28 countries by 2022.^6^

Evaluations indicate that RCs have a positive effect on students, including recovery-related domains such as wellbeing, empowerment, hope, and social inclusion.^7^ Previous evidence also suggests that RC attendance might lead to reduced health service use,^8^ including hospital admissions, involuntary admissions, and community contacts.^9,10^ However, these findings are based on relatively modest sample sizes, short follow-up periods, and the use of designs which are either un-controlled or in which students act as their own controls, i.e. without a control group of similar service users not attending the RC. There is clear need for more robust and rigorous evaluations to support causal inference.

This study was undertaken as part of the Recovery College Characterisation and Testing (RECOLLECT) 2 programme, which investigates the effectiveness and cost-effectiveness of RCs in England.^11^ Our aim was to conduct a large retrospective matched cohort study, in order to assess the impact of RCs on routinely collected outcomes and health service use over a considerable follow-up period. We therefore examined associations between RC enrolment and: mental health service use at an index mental health provider; use of other NHS hospital services for all causes; associated costs; and routinely collected outcomes among all students who enrolled at a RC in a diverse, urban area of South East London.

## Methods

The study protocol was published on the Open Science Framework in advance of the analysis (https://osf.io/6p3bs/). We created a retrospective matched cohort comprising mental health service user RC students and matched non-student service user controls from South London and Maudsley National Health Service (NHS) Foundation Trust (SLaM). SLaM provides comprehensive mental healthcare to an urban catchment comprising approximately 1·36 million people covering four London boroughs and is one of Europe’s largest specialist mental healthcare providers. The Clinical Record Interactive Search (CRIS) system is a platform and governance framework allowing access to de-identified SLaM records for research^12^. CRIS contains healthcare information for almost all people in contact with SLaM from January 1, 2007 and received research ethics committee approval as an anonymised data resource for secondary analyses (Oxford Research Ethics Committee C, reference 23/SC/0257).

To avoid the confounding effect of the COVID-19 pandemic on outcomes, the study end date was January 31, 2020, the date of the first recorded UK case of COVID-19. Therefore, the study period covers January 1, 2007 to January 31, 2020. The sample comprised students and controls with at least six months of follow-up data. For students, and their matched controls, the follow-up period began on the date the student enrolled at SLaM RC (the ‘index date’). Enrolment records at the SLaM RC covered the period March 13, 2013 (when SLaM RC registrations began) to December 31, 2019. Therefore, for a student to be included in the study they needed to have enrolled at the RC by July 31, 2019 to ensure at least six months follow-up data. Students and controls were followed up over three periods from the index date: six months, 12 months, and five years, allowing assessment of the short-term, mid-term and longer-term impact of RC enrolment, respectively.

People with lived experience of mental health challenges were involved in conceptualising and acquiring funding for the study, including as applicants, advisors and authors. We obtained advice from the RECOLLECT Lived Experience Advisory Panel, ensuring lived experience shaped the specific research question, and interpretation, and writing up of the findings.

## Procedure

We created the retrospective matched cohort in a two-stage data process:

### Stage 1: Control matching

Following data linkage and control identification (see Supplementary Methods), students and controls were matched based on several relevant sociodemographic and clinical variables listed later in the Methods. Only those with complete data on all matching variables were included in the cohort. Each control was then assigned the same RC enrolment date as their matched student. This ensured that both students and their corresponding controls shared the same study start date and follow-up period. Control service users who had not been matched with students were then removed from the dataset, resulting in the matched study cohort.

### Stage 2: Study outcome extraction

A dataset containing the matched cohort and their study start dates was returned to the CRIS team so that health service use and routinely collected HoNOS data could be added pre- and post-index date for each person in the cohort. Outcome data were collected, where applicable, across three post-enrolment timeframes: from the study start date to six months, 12 months, and five years. We also gathered outcome data for the equivalent pre-enrolment periods where applicable. All participants had six months of pre- and post-enrolment data available, while subsamples had data for the 12 month and five year periods.

## Measures

SLaM service use data included number of psychiatric inpatient admissions, associated bed days, and the number of involuntary admissions under the Mental Health Act (MHA). We examined the number of Home Treatment Team (HTT) and Community Mental Health Team (CMHT) events, and the number of liaison psychiatry attendances in general medical hospital emergency departments (EDs) in the SLaM catchment area. We also present ‘Active SLaM days’ showing the number of days students and controls were in an open episode of care during each follow-up period.

Other all-cause hospital use was measured in people who had data linkage with Hospital Episode Statistics (HES) available. To avoid double-counting, service use at SLaM was removed from the HES dataset prior to analysis (PROCODE=”RV5”). Outcomes included admitted patient care (finished consultant episode) and associated bed days. As well as overall hospital admissions, we examined elective and emergency admissions separately. We also assessed the numbers of ED and outpatient appointment attendances.

Routinely collected patient outcomes within SLaM services include the Health of the Nation Outcome Scales (HoNOS). The HoNOS is a 12-item staff-rated measure of health and social functioning in mental health services, with adequate psychometrics.^13^ Each item is scored on a 5-point scale (0=‘no problem’ to 4=‘severe problem’). The total score is the sum of all items, ranging from 0 (fewer or less severe difficulties) to 48 (very severe and wide-ranging problems). HoNOS was chosen as the primary measure because it was at the time the most widely collected outcome measure in routine practice both across England and in SLaM, maximising both the sample size for analysis and relevance to other services. Multiple HoNOS scores may be recorded for each patient but at irregular time points.

## Covariates and matching variables

Students and controls were matched on relevant sociodemographic and clinical variables. Sociodemographic variables included age at first face-to-face contact with SLaM services (caliper: five years), gender (exact match), and ethnicity (exact match) (Asian, Black (African, Caribbean, Other), Mixed, Other, Not stated/unknown, South Asian (Indian, Pakistani, Bangladeshi), White). Neighbourhood deprivation was measured using the Index of Multiple Deprivation (IMD) decile (caliper: two deciles), based on address at first face-to-face contact with SLaM. IMD scores are generated at the lower layer super output area (LSOA) level. LSOAs are geographical areas designed to support reporting of small area statistics and contain approximately 1 000 to 3 000 people. IMD scores are based on seven domains: income, employment, health and disability, education, skills and training, crime, barriers to housing and services, and living environment. The smoking status (exact match) of each student/control was determined based on whether they had a record of being a smoker at any time in the study period. Clinical variables used for matching included records of the most common mental health diagnoses amongst the students and controls based on ICD-10 block (exact match), including F2* (Schizophrenia, schizotypal and delusional disorders), F3* (Mood [affective] disorders), F4* (Neurotic, stress-related and somatoform disorders), and F6* (Disorders of adult personality and behaviour).

## Statistical analysis

Students and matched controls were compared on each characteristic using independent t-tests and χ2 tests to assess the success of the matching approach. Health service use data were positively skewed. Although medians and interquartile ranges (IQRs) would usually be reported in the case of skewed data, the median scores for almost all health service use data were zero due to the high number of students/controls who did not have any admissions or frequent contact with mental health services and other hospital services. Therefore, we also reported means and SDs for health service use outcomes.

For mental health and other all-cause hospital use, negative binomial regressions were used to handle skewed count data and to account for overdispersion (i.e. where the observed variance was greater than the expected variance). To assess the effect of being a student, interaction terms (intervention*time) were entered into models for each follow-up period. The coefficient of the interaction term represented the differential change in the student group relative to the control group, indicating the estimated effect of RC enrolment. Where exact matching could not be achieved for specific sociodemographic and clinical variables, these were accounted for through adjustment in the regression models.

To test the robustness of our controlled before-and-after findings relating to health service use, we performed a sensitivity analysis restricted to the student group only, i.e. an uncontrolled before-and-after study.

To model associated costs, mental health service use in the following categories was costed at the student/control level: number of bed days, HTT events, CMHT events, liaison psychiatry attendances in EDs, and admissions made under the MHA. Costs for all years were collected from the National Cost Collection 2023/24^14^ except for MHA admissions where a cost from 2020/21 had to be used and then inflated to 2023/24 prices. Costs from a single year were used to ensure consistent costs were applied and from the most recent cost year available. Unit costs and their descriptions are presented in Supplementary Material Table S1. Total mental healthcare costs were then calculated as the sum of these items for each patient. Inpatient admission costs were not included separately to avoid double counting, as the cost was assumed to be covered by the number of bed days. All costs are reported using 2023/24 prices. Costs were modelled using a Generalised Linear Model (GLM) with Gamma family and log link, to account for the right-skewed distribution of cost data. Models used the same time-treatment interaction as the main analysis and adjusted for the same covariates. To aid interpretation, we calculated average marginal effects, representing the difference in mean costs between treatment and control groups after adjusting for covariates. Other all-cause hospital use was costed and analysed using the same approach, with costs for hospital bed days, ED visits and outpatient appointments sourced from the National Cost Collection 2023/24^14^ (Table S1). To contextualise any observed cost reductions, we extrapolated findings to the national level where possible.

To determine the impact of RC enrolment on HoNOS scores, we applied a before-and-after design in students only, because the irregular collection of HoNOS meant control matching was not feasible. We included subsamples of students with completed HoNOS at each timepoint (i.e. six months, 12 months, five years) using the most recent score. For example, to assess the impact of RC enrolment on HoNOS scores at six months, we included students who had a HoNOS completed in the six-month period before enrolment, and in the six-month period after enrolment. We used negative binomial regression models due to the overdispersion of scores, and ordinal logistic regression to appropriately reflect the ordinal scale of each HoNOS item.

All analyses were performed using Stata 18·0 (StataCorp LLC, College Station, TX).

## Results Sample

Sample selection is depicted in the flow diagram in Figure 1. Our final sample comprised 1 435 students who had enrolled at the SLaM RC between March 13, 2013 and July 31, 2019, and 4 665 service user non-student controls (total N=6 100). Most students had four matched controls (62·9%, N=902), 13·8% (N=198) had three matched controls, 9·3% (N=134) had two, and 14·0% (N=201) had one matched control. The student and control samples are described in Table 1.

**Figure 1:**
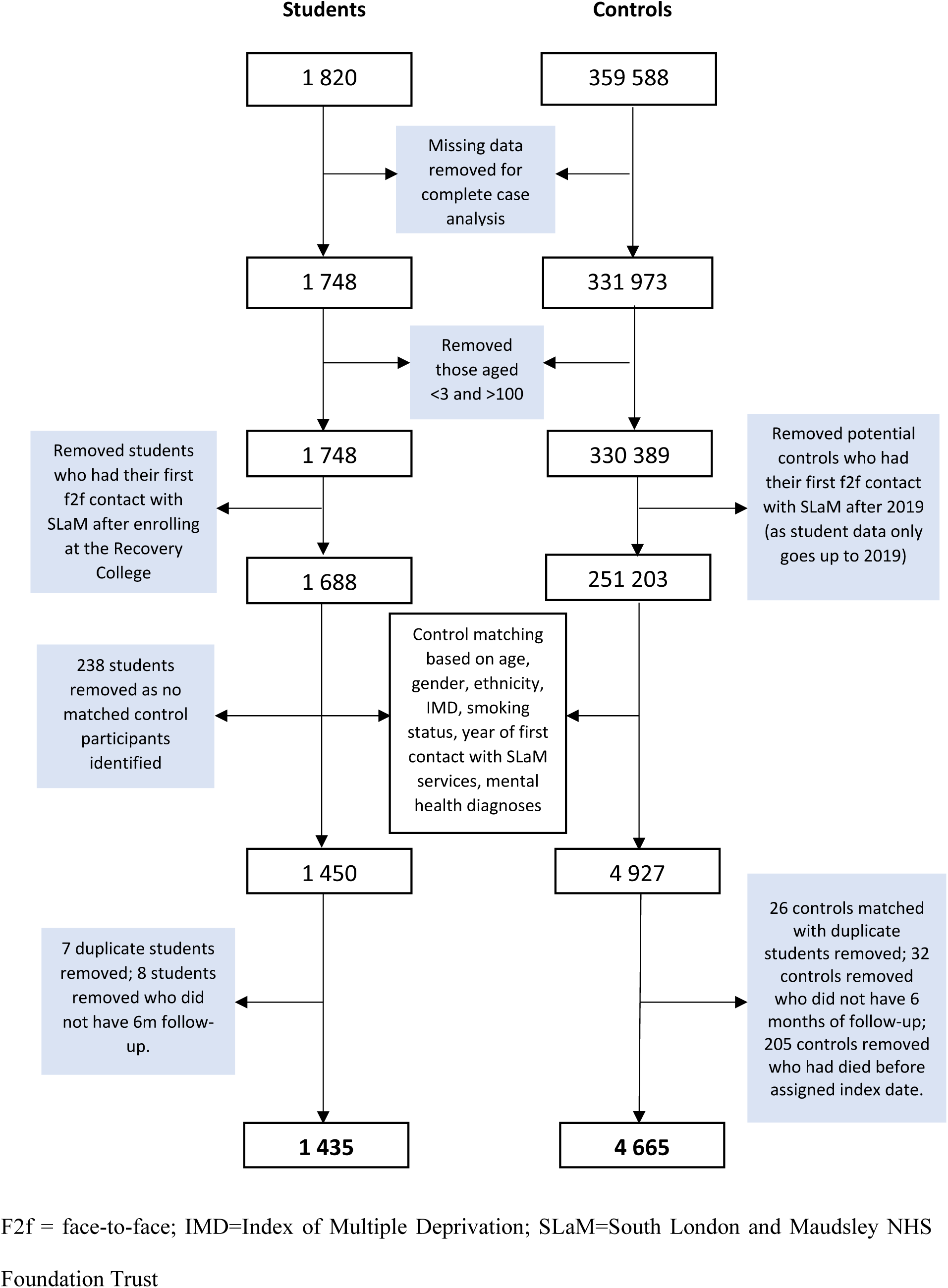
Sample selection flow chart.

**Table 1:** Sample characteristics for students and matched controls.

|  | Student (N=1435)<br>*M±SD or N(%)* | Control<br>*M±SD or N(%)* | P value |
| --- | --- | --- | --- |
| Age | 34·32±12·06 | 34·07±11·97 | 0·503 |
| Gender |  |  | 0·876 |
| *Male* | 579 (40·3) | 1893 (40·6) |  |
| *Female* | 856 (59·7) | 2772 (59·4) |  |
| Ethnicity |  |  | **0·039** |
| *White* | 801 (55·8) | 2703 (57·9) |  |
| *Mixed* | 47 (3·3) | 94 (2·0) |  |
| *South Asian* | 21 (1·5) | 52 (1·1) |  |
| *Black* | 391 (27·2) | 1286 (27·6) |  |
| *Other* | 120 (8·4) | 337 (7·2) |  |
| *Not stated* | 55 (3·8) | 193 (4·1) |  |
| Smoker | 868 (60·5) | 2670 (57·2) | **0·029** |
| IMD quintile |  |  | 0·170 |
| *1 (high deprivation)* | 363 (25·3) | 1108 (23·8) |  |
| *2* | 656 (45·7) | 2239 (48·0) |  |
| *3* | 293 (20·4) | 991 (21·2) |  |
| *4* | 96 (6·7) | 250 (5·4) |  |
| *5* | 27 (1·9) | 77 (1·6) |  |
| Diagnosis |  |  |  |
| *F2 ever* | 477 (33·2) | 1482 (31·8) | 0·296 |
| *F3 ever* | 673 (46·9) | 2086 (44·7) | 0·146 |
| *F4 ever* | 415 (28·9) | 1261 (27·0) | 0·161 |
| *F6 ever* | 322 (22·4) | 864 (18·5) | **0·001** |
| Years of first contact with<br>SLaM* |  |  | 0·998 |
| *2002* | 146 (10·2) | 469 (10·0) |  |
| Pre-index date HoNOS** | 10 (6 to 14)<br>*N=1280* | 9 (5 to 13)<br>*N=3082* | **<0·001** |

F2= Schizophrenia, schizotypal and delusional disorders; F3= Mood [affective] disorders; F4= Neurotic, stress-related and somatoform disorders; F6= Disorders of adult personality and behaviour; HoNOS=Health of the Nation Outcome Scale; IMD=Index of Multiple Deprivation; M=mean; SD=standard deviation
\* We conducted matching across 18 years (2001–2019). For brevity, we illustrate the process using 2002—the year with the highest student first contact rate—as an example.
\*\*Score closest to index date

Students and controls were well-matched in age, gender, F2* (schizophrenia, schizotypal and delusional disorders)/F3* (mood [affective] disorders) /F4* (neurotic, stress-related and somatoform disorders) diagnosis, IMD quintile and year of first contact with SLaM. Exact matching was not achieved for ethnicity, F6* (disorders of adult personality and behaviour) diagnosis, and smoking status. Regarding follow-up duration, 1 293 students and 4 197 controls had data available for the six and 12-month follow-up period, while 206 students and 670 controls were followed for five years.

## Impact of Recovery College enrolment on mental health service use and associated costs

Mental health service use across all follow-up periods is presented in Supplementary Table S2. At six months (student N=1 435, control N=4 665), reductions in inpatient admissions (aIRR=0·56, 95% CI=0·41 to 0·76), HTT events (aIRR=0·39, 95% CI=0·21 to 0·74), liaison psychiatry events (aIRR=0·43, 95% CI=0·30 to 0·64), and detentions under the MHA (aIRR=0·51, 95% CI=0·37 to 0·72) were significantly greater in those who enrolled at the RC than in the control group, as shown in Figure 2.

**Figure 2:**
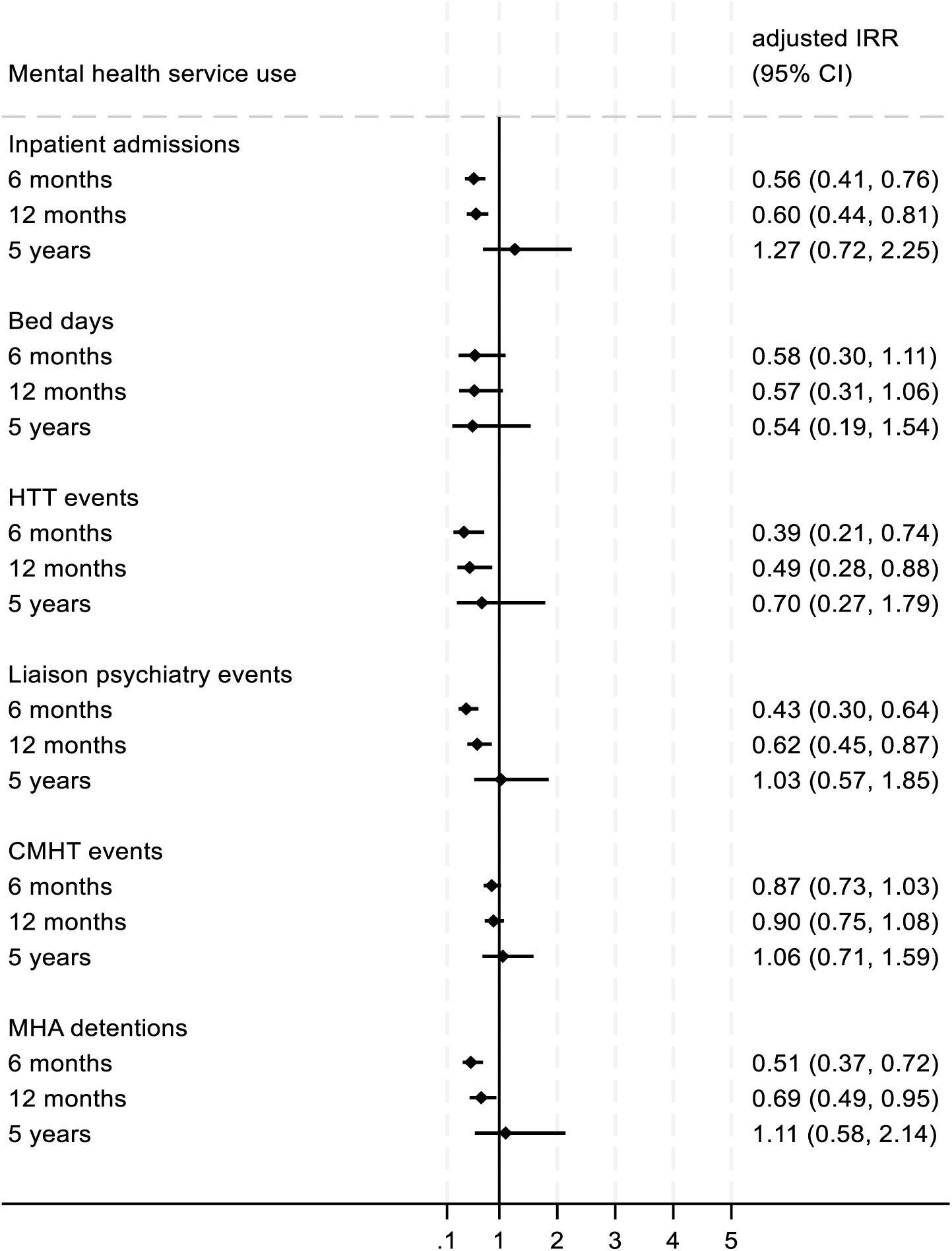
Associations between Recovery College enrolment and mental health service use at six months, 12 months, and five years. The adjusted incident rate ratios (aIRRs) shown represent the coefficients of the treatment*time interaction for each follow-up period. Lower aIRRs indicate greater reductions in mental health service use among the student group. For example, an aIRR of 0.56 corresponds to a 44% greater reduction in inpatient admissions at six months in students than in controls. Analyses were adjusted for ethnicity, smoking status, and F6* diagnosis. Sample sizes: 1435 students and 4665 controls at six months; 1293 students and 4197 controls at 12 months; 206 students and 670 controls at five years CI=confidence interval; CMHT=Community mental health team; HTT=Home treatment team; IRR=incident rate ratio; MHA=Mental Health Act.

At 12 months (student N=1 293, control N=4 197), significantly greater reductions in inpatient admissions, HTT events, liaison psychiatry events, and MHA detentions persisted. In those who had five years of follow-up (student N=206, control N=670) there were no significant differences in any mental health service use outcome.

Cost modelling is shown in Supplementary Table S3. Differences in the average marginal costs of mental health service use are shown in Figure 3 for the six month and 12-month follow-up periods.

**Figure 3:**
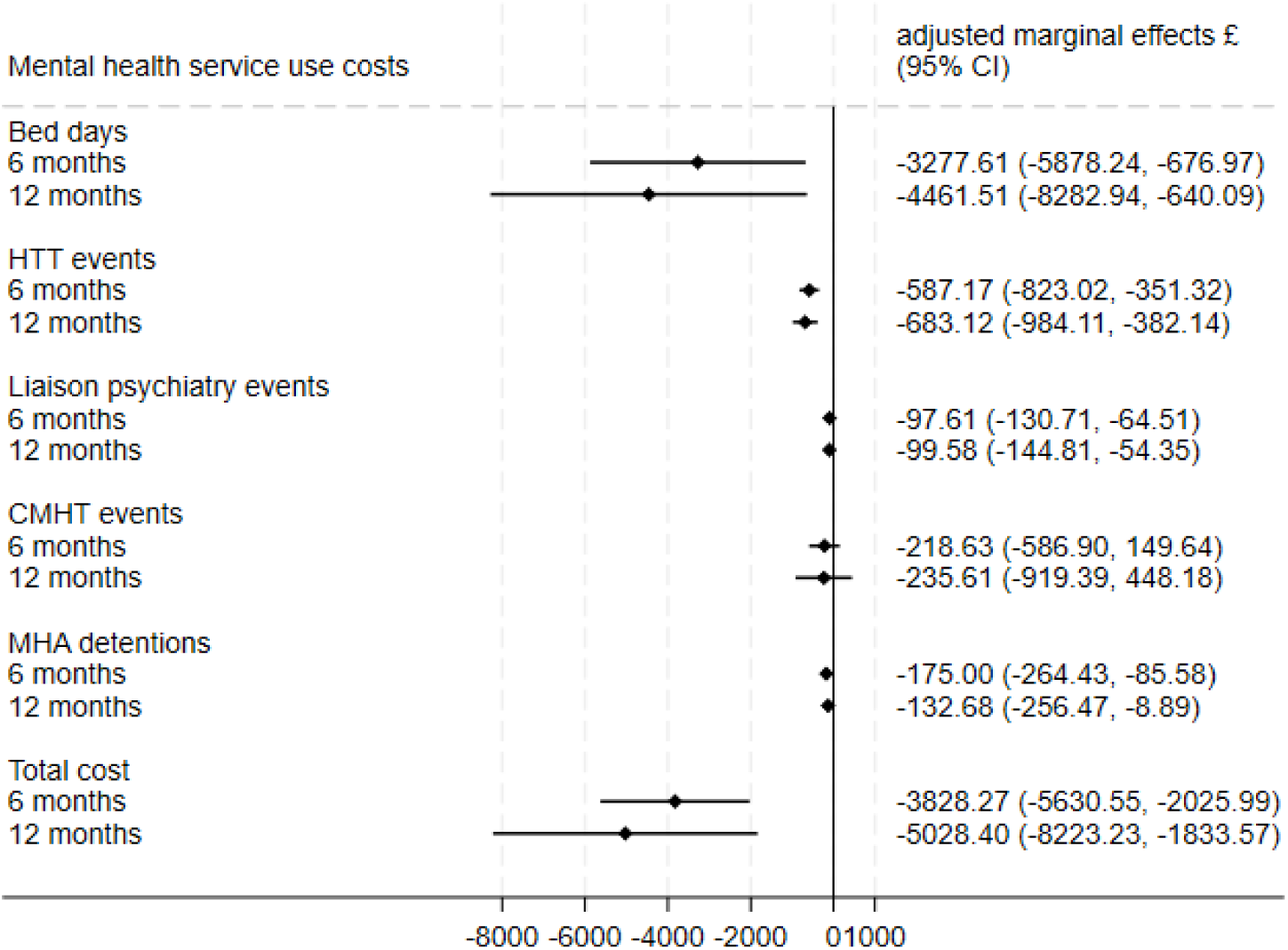
Adjusted marginal effects from GLM regression for mental health service use at six months and 12 months pre- to post-enrolment. The average marginal effects represent the difference in mean costs between treatment and control groups after adjusting for covariates. Analyses were adjusted for ethnicity, smoking status, and F6* diagnosis. GLM = Generalised Linear Model; CMHT = Community Mental Health Team; HTT = home Treatment Team; MHA=Mental Health Act.

At six months, students showed significantly greater cost reductions than controls related to bed days, HTT contacts, liaison psychiatry interventions, and detentions under the MHA. This was reflected in a significantly larger reduction in total costs per student compared with controls (£3 828 (95% CI -£5 631 to -£2 025)). By 12 months, the reduction in total costs among students was even more pronounced relative to controls (£5 028 (95% CI -£8 223 to -£1 834)).

### Impact of Recovery College enrolment on other all-cause hospital use and associated costs

Linked Hospital Episode Statistics (HES) were available for 4 701 people (Student N=1 193, Control N=3 508). Other all-cause hospital use is presented in Table S4. At six months, significantly greater reductions in hospital bed days (aIRR=0·53, 95% CI=0·35 to 0·81) and ED attendances (aIRR=0·74, 95% CI=0·60 to 0·91) were seen in students relative to controls, see Figure 4.

**Figure 4:**
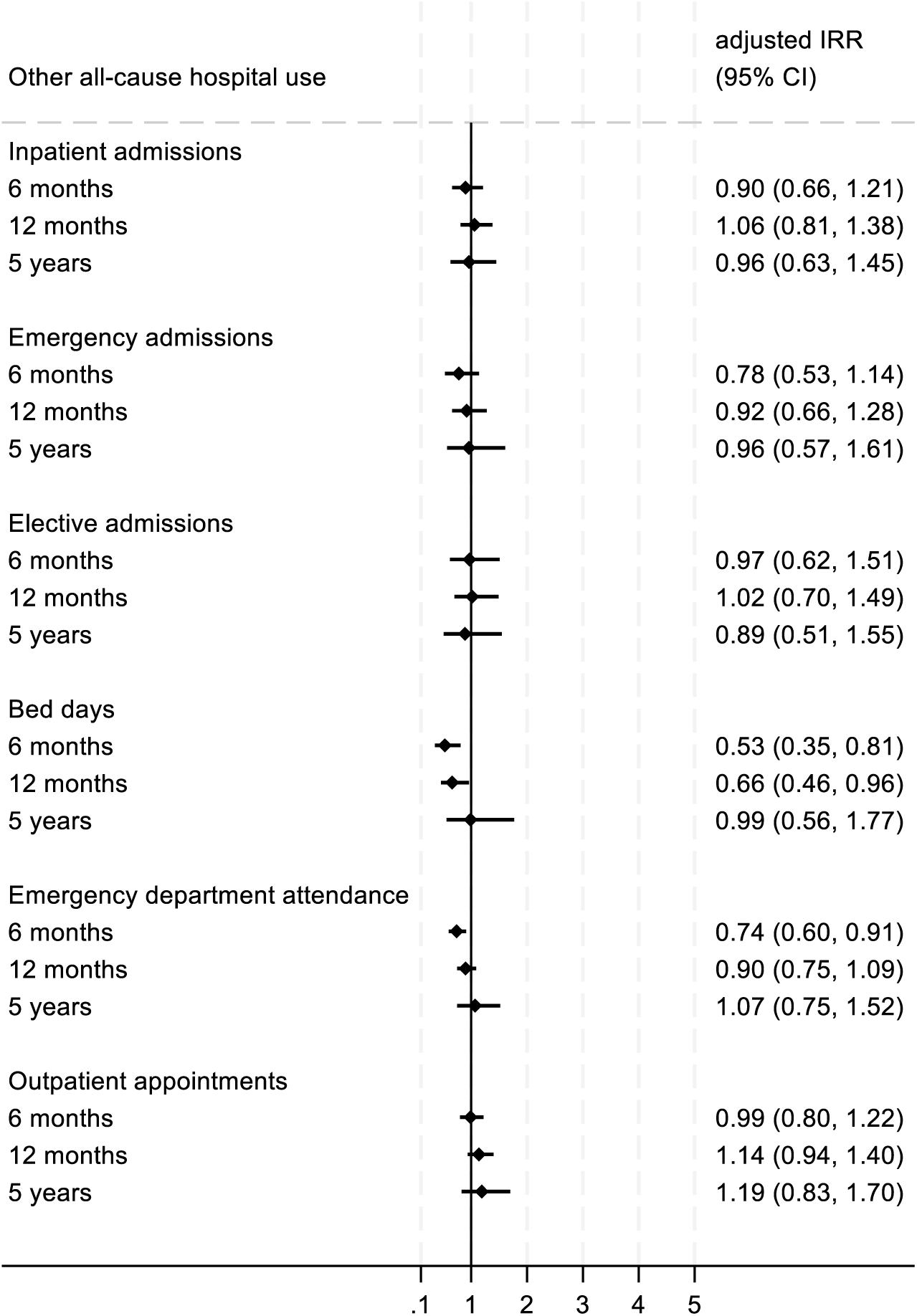
Associations between Recovery College enrolment and other all-cause hospital use at six months, 12 months, and five years. The adjusted incident rate ratios (aIRRs) shown represent the coefficients of the treatment*time interaction for each follow-up period. Lower aIRRs indicate greater reductions in other hospital use among the student group. For example, an aIRR of 0.53 corresponds to a 47% greater reduction in bed days at six months in students than in controls. Analyses were adjusted for F6* diagnosis. Sample sizes: 1193 students and 3508 controls at six months; 1072 students and 3158 controls at 12 months; 180 students and 529 controls at five years. CI=confidence interval; IRR=incident rate ratio

Greater reductions in hospital bed days persisted at 12 months post-enrolment (student N=1 072, control N=3 158) (aIRR=0·66, 95% CI=0·46 to 0·96) but reductions in ED attendances were no longer significant. At five years (student N=180, control N=529), there were no significant differences in general health service use.

Results for costs analysis of other all-cause hospital use are reported in Supplementary Table S5. At six months, total costs were £443 (95% CI -£792 to -£94) lower per student than per control, primarily due to reductions in bed days. Differences in costs at 12 months and five years were not statistically significant.

### Cost extrapolation

Previously we have reported that in 2021, there were 88 RCs operating in England offering 11 000 courses to approximately 45 000 students, with an estimated NHS spend of £13·4 million.^15^ Drawing on a three-college casemix study which found that 67.5% of students were also service users,^16^ we conservatively assumed that 50% of patients nationally were service users. Applying the observed mean reduction in serviced use costs (£5 000 per patient), we estimated potential national savings. Based on these assumptions, the extrapolation suggests potential cost savings of £112·5m (22 500 × £5 000), equivalent to an 8·4:1 financial return on investment.

### The impact of Recovery College enrolment on HoNOS

A before-and-after analysis in the student group demonstrated a significant decrease in total HoNOS scores across all timepoints, shown in Table 2.

**Table 2:** Impact of Recovery College enrolment on HoNOS total scores.

|  | **Student**<br>***N*** | **Pre-index**<br>**date**<br>***Median***<br>***(IQR)*** | **Post-index**<br>**date**<br>***Median***<br>***(IQR)*** | **Change over time**<br>***IRR (95% CI)*** |
| --- | --- | --- | --- | --- |
| **Six months** | 475 | 10 (7 to 14) | 9 (6 to 13) | *0.90 (0.84 to 0.96) |
| **12 months** | 793 | 10 (7 to 14) | 9 (6 to 14) | *0.92 (0.87 to 0.97) |
| **Five years** | 979 | 10 (7 to 14) | 9 (6 to 14) | *0.93 (0.88 to 0.98) |

CI=confidence interval; HoNOS=Health of the Nation Outcome Scale; IQR=interquartile range; IRR=incident rate ratio. \* $p < 0.05$ , \*\* $p < 0.001$

At six months post-enrolment, there was a significant 10% reduction in HoNOS total score compared with pre-enrolment (N=475; IRR=0·90, 95% CI=0·84 to 0·96). Median scores for individual items for each follow-up period and results from negative binomial and ordinal regression models are presented in Supplementary Table S6. There were significant reductions in agitated behaviour (OR=0·70, 95% CI=0·55 to 0·91), hallucinations (OR=0·75, 95% CI=0·59 to 0·97), depressed mood (OR=0·74, 95% CI=0·59 to 0·93), and ‘other mental problems’ (OR=0·76, 95% CI=0·60 to 0·95). At both 12 months (N=793) and five years (N=979), 8% and 7% reductions in total HoNOS scores were seen respectively, and this reduction was driven by similar HoNOS items as at six months. Additionally, self-injury and problem with occupation scores showed significant reductions at 12 months, while an increase in physical illness was noted at five years.

## Sensitivity analyses

Results from an uncontrolled before-and-after study performed on the student sample relating to health service use are presented in Tables S7 and S8. Before-and-after analyses produced similar results to controlled models.

## Discussion

This is the largest study of its kind, employing a methodologically rigorous approach to evaluate the impact of Recovery Colleges on service use, costs, and service user outcomes. In comparison with controls, RC enrolment was linked to significant reductions in both mental health service use at an index mental health provider and wider all-cause hospital use nationally at six- and 12-months post-enrolment. These reductions translated into cost savings of approximately £5 000 for mental health services at 12 months and £400 for general health services at six months. These findings indicate an 8·4:1 financial return on investment. These effects were not sustained at five years, which may indicate that effects do not persist but may also reflect insufficient statistical power to detect longer-term impacts. In a subsample of students with available data, RC enrolment was associated with significant improvements in health and social functioning, which was maintained up to five years.

These positive findings align with, and considerably strengthen, the findings of other evaluative reviews of RCs.^9,10^ They also confirm a component of the RECOLLECT change model,^17^ namely changes in health service use as an outcome for students. Qualitative studies suggest that both the nature of the relationships established at RCs and what is learned on courses are likely to play a role, empowering students to engage differently with health services and/or with other people and organisations in their lives and to self-manage their illness.^7^

A 2025 review identified the need for studies with adequate statistical power,^7^ which we were able to address for the 6 and 12 month periods through a large sample size (6 100 in total), and appropriate control group matching on several sociodemographic and clinical characteristics, including deprivation, which has until now been largely overlooked. Other key strengths of this study include the use of linked electronic records, enabling health service use and outcome assessment over a five-year period in the largest sample of RC students to date (n=1 435). The complete case ascertainment and controlled before-and-after design enhanced methodological rigour, minimised bias, and supported robust inferences about the impact of RC enrolment.

Several limitations apply. Although students and controls were matched on several relevant sociodemographic and clinical variables, we cannot rule out unmeasured confounding (e.g. lifestyle factors, social support). We were also unable to achieve exact matching on certain characteristics (e.g. ethnicity, smoking status) indicating inherent differences between student and controls. Moreover, RC students appeared to use mental health services more frequently than matched controls in both pre- and post-enrolment periods, further suggesting underlying differences between the groups that were not fully captured. To account for this, we focused on changes in resource use and outcomes over time between students and controls rather than direct group comparisons and conducted an uncontrolled before-and-after analysis within students alone to test the robustness of our findings. Due to how student data is collected, it was only possible to assess the impact of RC enrolment; a significant minority of people who enrol do not attend any courses. Future studies should consider assessing how length of attendance, number of courses undertaken, and type of course^18^ impact health service use. To enable this, RCs should consider collecting this information routinely. Inconsistent collection of HoNOS limited our sample size. The single site design limits generalisability of the findings which are primarily applicable to service user students in diverse urban settings. Future studies might also evaluate the wider impact of RC enrolment, including primary care, social care and adult education services. Finally, our original study design was a five-site study,^11^ but for reasons we will report elsewhere (e.g. data availability, research capacity), only one site proved possible.

We identify three avenues of future research. First, understanding the mechanisms of action of RCs remains a priority. RECOLLECT is based on a multi-level change model,^11^ which identifies student-, service- and societal-level outcomes. Important under-researched elements include social inclusion outcomes,^19^ how RC co-production influences peer-practitioner relationships,^20^ and societal outcomes such as occupation and impact on family members.^21^ The RECOLLECT Change Model will be refined in the light of findings from this and other studies of RCs for general^22^ and specific^23^ populations, including post-COVID practice changes^24^ and online colleges.^25^

Second, our study demonstrates that rigorous research with large samples and appropriate statistical analysis is possible with RCs, without the need for randomisation. In England, RCs are so widely available that clinical equipoise does not exist meaning randomisation would be ethically problematic; therefore, non-RCT designs are more suitable. Some researchers agree with this,^22^ whereas others identify the need for crossover designs and RCTs.^7^ To our knowledge, no RCT of RCs has reported or is currently underway.

Finally, RCs are spreading globally. We published a survey identifying 221 colleges in 28 countries in 2022,^6^ and we are informally aware of 274 colleges operating in 31 countries in 2025. Established cultural influences on fidelity include individualism, indulgence, uncertainty avoidance and long-term orientation.^26^ These impact operational aspects including equality, learning focus, co-production, and community focus.^27^ Therefore, the impact of RC attendance beyond England needs investigation with equivalent methodological rigour, using co-created methods^28^ to identify invariant versus culturally-influenced mechanisms.

In conclusion, our study evidence supports ongoing investment in RCs with significant return on investment, especially in England. It aligns with existing calls to scale up RC provision,^29^ and strengthens the evidence of benefits for service users, services and society.^30^

## Data Availability

The data for this study were obtained via the Clinical Record Interactive Search (CRIS) system. CRIS provides researchers affiliated with the National Institute for Health Research (NIHR) Maudsley Biomedical Research Centre (BRC) secure access to de-identified electronic health records for approved research projects. Applications for access to de-identified data can be submitted through the NIHR BRC at the South London and Maudsley NHS Foundation Trust. Because the source data are derived from patient records, access is restricted to eligible researchers who have obtained the necessary ethical and institutional approvals. All requests are subject to review and authorization by the CRIS Oversight Committee. Further information about CRIS and its data access policies is available at: https://www.maudsleybrc.nihr.ac.uk/facilities/clinical-record-interactive-search-cris/.

## Contributors

AR, TA, IB, RE, TJ, DH, MN, SM, RAE, CH and MS conceptualised the study. DD, HH-B, SKT, MM, JS, under the supervision of TJ, DH, IB, RE, MN, RAE, CH and MS, were responsible for data collection. AR, TA, IB, RE, CH and MS were responsible for data analysis. All authors were involved in data interpretation. AR, TA, RAE, CH and MS were involved in the writing the original draft and are responsible for the decision to submit the manuscript. All authors were involved in reviewing and editing the manuscript and approved the final version. All authors had access to all data, and AR, TA, CH and MS have accessed and verified the data.

## Acknowledgments

This article is independent research funded by the NIHR (Programme Grants for Applied Research, Recovery Colleges Characterisation and Testing (RECOLLECT) 2, NIHR200605) and part funded by the NIHR Maudsley Biomedical Research Centre at South London and Maudsley NHS Foundation Trust and King’s College London. The views expressed are those of the authors and not necessarily those of the NIHR or the Department of Health and Social Care. We would like to acknowledge NHS England as the provider of the Hospital Episode Statistics (HES) data; those who carried out the original collection and analysis of the data bear no responsibility for their further analysis or interpretation. We thank the RECOLLECT Lived Experience Advisory Panel for their many contributions. MS acknowledges the support of NIHR Nottingham Biomedical Research Centre.

