## Supplementary Material for "The impact of Recovery College enrolment on health service use and patient outcomes: retrospective matched cohort study using routinely collected data"

**Table of contents**

| Page | Contents |
|---|---|
| 2 | Supplementary Methods |
| 3 | Table S1: Unit costs for health care resource use |
| 4 | Table S2: Mental health service use in pre- and post-index date periods presented as both medians and IQRs and means and SDs alongside results from unadjusted and adjusted negative binomial regression models |
| 6 | Table S3: Impact of Recovery College enrolment on mental health service costs |
| 8 | Table S4: Other all-cause hospital use in pre- and post-index date periods presented as both medians and IQRs and means and SDs alongside results from unadjusted and adjusted negative binomial regression models |
| 10 | Table S5: Impact of Recovery College enrolment on other all-cause hospital use costs |
| 11 | Table S6: Impact of Recovery College enrolment on HoNOS scores: Before-and-after study in students only |
| 12 | Table S7: Impact of Recovery College enrolment on mental health service use: Before-and-after study in student group |
| 13 | Table S8: Impact of Recovery College enrolment on other all-cause hospital use: Before-and-after study in student group |

### Supplementary Methods

*Control matching*

Student enrolment details (name, date of birth, Recovery College enrolment date) were sent securely from SLaM Recovery College to the National Institute for Health and Care Research (NIHR) Biomedical Research Centre (BRC) at SLaM who manage the CRIS system. Students were identified within CRIS so that the research team could be provided with an anonymised dataset comprising SLaM Recovery College students and all SLaM service users who were 16 years or older at their first attended contact with SLaM services. This dataset contained sociodemographic and clinical variables and was used for control matching. Controls were identified using 1:4 caliper matching without replacement (STATA module '*calipmatch'* https://github.com/michaelstepner/calipmatch). The search for matched controls was performed 'greedily', meaning it was possible that some students would end up either unmatched or with less than four matches because all possible matching controls had already been matched with another case.

### Table S1: Unit costs for health care resource use

| **Mental Health resource use from SLAM** | | | |
|---|---|---|---|
| **Resource use variable collected** | **Cost description** | **Source** | **Unit cost** |
| Bed days | Acute Mental Health Care [code: 10MHPS] | National Cost Collection 2023/24 | £575 |
| Home Treatment Team events | Crisis Resolution Team/Home Treatment Service [code: A02] | National Cost Collection 2023/24 | £338 |
| Liaison psychiatry in emergency departments | Psychiatric Liaison Service [code: A11] | National Cost Collection 2023/24 | £355 |
| Community Mental Health Team events | Community Mental Health Team – Functional [code: A06] | National Cost Collection 2023/24 | £280 |
| Mental Health Act detention | Mental Health Care Cluster Initial Assessment – Psychotic Crisis [code: MHCC14] | National Cost Collection 2020/21 | £612* |
| | Ambulance – see & convey [code: 04] | National Cost Collection 2023/24 | £459 |
| **General hospital resource use from HES** | | | |
| Bed days | Weighted average of Admitted Patient Care, Regular Day or Night Admissions | National Cost Collection 2023/24 | £400 |
| A&E attendances | Weight average of Emergency Care, all services | National Cost Collection 2023/24 | £273 |
| Outpatient appointments | Weight average of Outpatient Care, all services | National Cost Collection 2023/24 | £176 |

A&E=Accident& Emergency; HES=Hospital Episode Statistics; SLaM=South London and Maudsley NHS Foundation Trust
*unit cost £533 in 2020/21 prices, inflated to 2023/24 using PSSRU 2024 [https://kar.kent.ac.uk/109563/]

**Table S2: Mental health service use in pre- and post-index date periods presented as both medians and IQRs and means and SDs alongside results from unadjusted and adjusted negative binomial regression models**

| | | | | | | |
|---|---|---|---|---|---|---|
| **6 months** | Student (N=1 435) | | Control (N=4 665) | | | |
| | 6m pre | 6m post | 6m pre | 6m post | | |
| | *Median (IQR)* | *Median (IQR)* | *Median (IQR)* | *Median (IQR)* | | |
| Active SLaM days | 181 (88 to 184) | 182 (86 to 184) | 0 (0 to 181) | 0 (0 to 182) | | |
| Inpatient admissions | 0 (0 to 0) | 0 (0 to 0) | 0 (0 to 0) | 0 (0 to 0) | | |
| All bed days | 0 (0 to 0) | 0 (0 to 0) | 0 (0 to 0) | 0 (0 to 0) | | |
| HTT events | 0 (0 to 0) | 0 (0 to 0) | 0 (0 to 0) | 0 (0 to 0) | | |
| Liaison events | 0 (0 to 0) | 0 (0 to 0) | 0 (0 to 0) | 0 (0 to 0) | | |
| CMHT events | 7 (2 to 14) | 6 (1 to 14) | 0 (0 to 2) | 0 (0 to 2) | | |
| MHA detentions | 0 (0 to 0) | 0 (0 to 0) | 0 (0 to 0) | 0 (0 to 0) | | |
| | *Mean (SD)* | *Mean (SD)* | *Mean (SD)* | *Mean (SD)* | *Unadjusted IRR (95% CI)* | *Adjusted IRR (95% CI)* |
| Active SLaM days | 136·65 (67·76) | 136·84 (72·44) | 63·45 (81·54) | 62·90 (82·38) | | |
| Inpatient admissions | 0·35 (0·85) | 0·19 (0·65) | 0·09 (0·47) | 0·09 (0·45) | 0·60 (0·44 to 0·83)* | 0·56 (0·41 to 0·76)** |
| All bed days | 13·95 (38·64) | 9·26 (34·10) | 3·03 (17·46) | 2·78 (16·83) | 0·73 (0·36 to 1·44) | 0·58 (0·30 to 1·11) |
| HTT events | 2·45 (8·48) | 1·23 (5·89) | 0·46 (3·25) | 0·45 (3·21) | 0·51 (0·27 to 0·98)* | 0·39 (0·21 to 0·74)* |
| Liaison events | 0·43 (1·46) | 0·21 (1·06) | 0·12 (0·68) | 0·11 (0·68) | 0·55 (0·37 to 0·82)* | 0·43 (0·30 to 0·64)** |
| CMHT events | 10·21 (13·37) | 10·08 (15·22) | 2·74 (6·22) | 2·96 (7·58) | 0·91 (0·76 to 1·09) | 0·87 (0·73 to 1·03) |
| MHA detentions | 0·26 (0·77) | 0·16 (0·96) | 0·08 (0·41) | 0·08 (0·40) | 0·59 (0·42 to 0·85)* | 0·51 (0·37 to 0·72)** |
| **12 months** | Student (N=1 293) | | Control (N=4 197) | | | |
| | 12m pre | 12m post | 12m pre | 12m post | | |
| | *Median (IQR)* | *Median (IQR)* | *Median (IQR)* | *Median (IQR)* | | |
| Active SLaM days | 341 (125 to 365) | 366 (126 to 366) | 4 (0 to 305) | 0 (0 to 323) | | |
| Inpatient admissions | 0 (0 to 0) | 0 (0 to 0) | 0 (0 to 0) | 0 (0 to 0) | | |
| All bed days | 0 (0 to 0) | 0 (0 to 0) | 0 (0 to 0) | 0 (0 to 0) | | |
| HTT events | 0 (0 to 0) | 0 (0 to 0) | 0 (0 to 0) | 0 (0 to 0) | | |
| Liaison events | 0 (0 to 0) | 0 (0 to 0) | 0 (0 to 0) | 0 (0 to 0) | | |
| CMHT events | 11 (3 to 26) | 10 (2 to 27) | 0 (0 to 5) | 0 (0 to 5) | | |
| MHA detentions | 0 (0 to 0) | 0 (0 to 0) | 0 (0 to 0) | 0 (0 to 0) | | |
| | *Mean (SD)* | *Mean (SD)* | *Mean (SD)* | *Mean (SD)* | *Unadjusted IRR (95% CI)* | *Adjusted IRR (95% CI)* |
| Active SLaM days | 250·19 (136·86) | 258·78 (145·01) | 123·43 (154·07) | 120·12 (157·3) | | |
| Inpatient admissions | 0·52 (1·26) | 0·30 (0·96) | 0·15 (0·66) | 0·13 (0·62) | 0·66 (0·49 to 0·91)* | 0·60 (0·44 to 0·81)* |
| All bed days | 23·21 (66·57) | 16·65 (60·08) | 5·43 (29·70) | 4·90 (28·51) | 0·79 (0·42 to 1·51) | 0·57 (0·31 to 1·06) |
| HTT events | 3·44 (10·10) | 1·99 (8·18) | 0·87 (5·05) | 0·78 (4·48) | 0·64 (0·35 to 1·15) | 0·49 (0·28 to 0·88)* |
| Liaison events | 0·65 (1·90) | 0·39 (1·62) | 0·23 (0·98) | 0·19 (1·02) | 0·72 (0·51 to 1·02) | 0·62 (0·45 to 0·87)* |
| CMHT events | 17·90 (23·41) | 18·32 (26·29) | 5·35 (11·41) | 5·54 (12·72) | 0·99 (0·82 to 1·19) | 0·90 (0·75 to 1·08) |
| MHA detentions | 0·35 (1·03) | 0·26 (1·34) | 0·12 (0·52) | 0·11 (0·50) | 0·75 (0·53 to 1·06) | 0·69 (0·49 to 0·95)* |
| **5 years** | Student (N=206) | | Control (N=670) | | | |
| | 5y pre | 5y post | 5y pre | 5y post | | |
| | *Median (IQR)* | *Median (IQR)* | *Median (IQR)* | *Median (IQR)* | | |
| Active SLaM days | 1075 (352 to 1775) | 937 (372 to 1827) | 429·5 (54 to 1173) | 154 (0 to 1016) | | |
| Inpatient admissions | 0 (0 to 1) | 0 (0 to 0) | 0 (0 to 1) | 0 (0 to 0) | | |
| All bed days | 0 (0 to 24) | 0 (0 to 0) | 0 (0 to 3) | 0 (0 to 0) | | |
| HTT events | 0 (0 to 9) | 0 (0 to 0) | 0 (0 to 0) | 0 (0 to 0) | | |
| Liaison events | 0 (0 to 1) | 0 (0 to 1) | 0 (0 to 1) | 0 (0 to 0) | | |

| | | | | | | |
|---|---|---|---|---|---|---|
| CMHT events | 41·5 (10 to 94) | 34 (9 to 84) | 8 (1 to 42) | 2 (0 to 24) | | |
| MHA detentions | 0 (0 to 0) | 0 (0 to 0) | 0 (0 to 0) | 0 (0 to 0) | | |
| | *Mean (SD)* | *Mean (SD)* | *Mean (SD)* | *Mean (SD)* | *Unadjusted IRR (95% CI)* | *Adjusted IRR (95% CI)* |
| Active SLaM days | 1030·70 (662·09) | 988·79 (668·45) | 657·58 (654·99) | 546·13 (686·59) | | |
| Inpatient admissions | 1·43 (3·23) | 0·98 (2·92) | 0·94 (2·58) | 0·48 (1·74) | 1·35 (0·72 to 2·56) | 1·27 (0·72 to 2·25) |
| Bed days | 62·00 (209·24) | 36·31 (141·85) | 25·26 (86·08) | 19·21 (86·77) | 0·77 (0·26 to 2·28) | 0·54 (0·19 to 1·54) |
| HTT events | 9·09 (19·84) | 6·13 (17·73) | 4·83 (17·30) | 3·61 (14·77) | 0·90 (0·34 to 2·42) | 0·70 (0·27 to 1·79) |
| Liaison psychiatry | 1·49 (3·59) | 1·32 (3·93) | 1·09 (3·58) | 0·84 (4·12) | 1·14 (0·61 to 2·14) | 1·03 (0·57 to 1·85) |
| CMHT events | 71·81 (108·00) | 58·86 (74·69) | 30·82 (53·80) | 25·95 (54·79) | 0·97 (0·64 to 1·48) | 1·06 (0·71 to 1·59) |
| MHA detentions | 0·77 (2·16) | 0·62 (1·92) | 0·57 (1·69) | 0·35 (1·28) | 1·29 (0·62 to 2·70) | 1·11 (0·58 to 2·14) |

CI=confidence interval; CMHT=Community Mental Health Teams; HTT=Home Treatment Team; IQR=interquartile range; IRR=incident rate ratio; MHA=Mental Health Act; SD=standard deviation; SLaM=South London and Maudsley NHS Foundation Trust
*p<0·05, **p<0·001
The IRRs presented are coefficients of the interaction term – treatment*time – for each follow-up period· Adjusted models included ethnicity, smoking status, and F6 diagnosis·

| Table S3: Impact of Recovery College enrolment on mental health service costs | | | | | | | |
|---|---|---|---|---|---|---|---|
| | Student N=1 435 | | Control N=4 665 | | | | |
| | *Mean (SD)* | | *Mean (SD)* | | | | |
| *Costs (£)* | *6 months pre* | *6 months post* | *6 months pre* | *6 months post* | *Unadjusted IRR (95% CI)* | *Adjusted IRR (95% CI)* | *Marginal Effects £ (95% CI)* |
| Bed days | 7990·23 (22168·78) | 5322·04 (19567·42) | 1733·2 (10005·8) | 1586·03 (9644·47) | 0·73 (0·52 to 1·02) | 0·56 (0·35 to 0·90)* | -3277·61 (-5878·24 to -676·97)* |
| HTT events | 830·13 (2862·33) | 426·07 (2005·21) | 153·89 (1094·3) | 150·72 (1081·64) | 0·51 (0·34 to 0·78)** | 0·38 (0·24 to 0·61)** | -587·17 (-823·02 to -351·32)** |
| Liaison psychiatry | 158·93 (548·37) | 77·25 (377·39) | 43·85 (239·47) | 39·08 (240·79) | 0·55 (0·38 to 0·82)** | 0·41 (0·28 to 0·59)** | -97·61 (-130·71 to -64·51)** |
| CMHT events | 2857·44 (3751·58) | 2823·87 (4259·72) | 765·41 (1740·03) | 824·97 (2116·38) | 0·91 (0·79 to 1·05) | 0·87 (0·74 to 1·01) | -218·63 (-586·90 to 149·64) |
| MHA detentions | 275·36 (826·91) | 170·71 (1028·31) | 82·79 (433·43) | 85·75 (422·86) | 0·60 (0·40 to 0·89)* | 0·40 (0·24 to 0·67)** | -175·00 (-264·43 to -85·58)** |
| Total | 12112·07 (23587·49) | 8819·93 (20868·63) | 2779·13 (11099·66) | 2686·55 (10864·24) | 0·75 (0·60 to 0·95)* | 0·66 (0·52 to 0·83)* | -3828·27 (-5630·55 to -2025·99)* |
| | Student N=1 293 | | Control N=4 197 | | | | |
| | *Mean (SD)* | | *Mean (SD)* | | | | |
| *Costs (£)* | *12 months pre* | *12 months post* | *12 months pre* | *12 months post* | *Unadjusted IRR (95% CI)* | *Adjusted IRR (95% CI)* | *Marginal Effects £ (95% CI)* |
| Bed days | 12964 (37428·02) | 8949·76 (33063·67) | 3103·48 (16973·74) | 2705·17 (15829·7) | 0·80 (0·56 to 1·13) | 0·56 (0·36 to 0·87)* | -4461·51 (-8282·94 to -640·09)* |
| HTT events | 1133·46 (3309·55) | 665·69 (2703·69) | 282·51 (1650·56) | 253·86 (1486·91) | 0·64 (0·44 to 0·92)* | 0·49 (0·33 to 0·74)* | -683·12 (-984·11 to -382·14)* |
| Liaison psychiatry | 246·51 (783·06) | 138·26 (566·89) | 79·38 (342·23) | 67·51 (368·44) | 0·73(0·51 to 1·03) | 0·60 (0·43 to 0·84)* | -99·58 (-144·81 to -54·35)* |
| CMHT events | 4995·95 (6461·07) | 4947·25 (7113·11) | 1471·85 (3144·78) | 1512·69 (3485·02) | 0·99 (0·86 to 1·14) | 0·90 (0·77 to 1·05) | -235·61 (-919·39 to 448·18) |
| MHA detentions | 376·3 (1091·18) | 259·77 (1374·51) | 126·58 (573·32) | 121·56 (539·76) | 0·75 (0·51 to 1·10) | 0·64 (0·40 to 1·02) | -132·68 (-256·47 to -8·89) |
| Total | 19716·22 (39581·51) | 14960·73 (34964·61) | 5063·79 (18736·86) | 4660·79 (17688·04) | 0·83 (0·66 to 1·05) | 0·70 (0·56 to 0·88)* | -5028·40 (-8223·23 to -1833·57)* |
| | Student N=206 | | Control N=670 | | | | |
| | *Mean (SD)* | | *Mean (SD)* | | | | |
| *Costs (£)* | *5 years pre* | *5 years post* | *5 years pre* | *5 years post* | *Unadjusted IRR (95% CI)* | *Adjusted IRR (95% CI)* | *Marginal Effects £ (95% CI)* |
| Bed days | 36337·37 (107749·98) | 18573·74 (74166·2) | 15986·37 (64921·71) | 7915·56 (40726·77) | 0·77 (0·34 to 1·75) | 0·54 (0·20 to 1·47) | -11207·53 (-32732·37 to 10317·31) |
| HTT events | 2822·52 (6549·97) | 1665·17 (5456·19) | 1309·32 (5116·84) | 698·67 (3265·58) | 0·90 (0·47 to 1·72) | 0·69 (0·31 to 1·53) | -1439·75 (-3650·74 to 771·25) |
| Liaison psychiatry | 636·69 (1935·28) | 389·44 (1726·08) | 297·77 (955·53) | 204·56 (917·23) | 1·14 (0·58 to 2·27) | 1·00 (0·54 to 1·84) | -4·22 (-251·55 to 243·12) |
| CMHT events | 16116·78 (22409) | 11540·93 (15419·12) | 6886·77 (12739·16) | 4335·17 (9923·65) | 0·97 (0·69 to 1·37) | 1·07 (0·74 to 1·54) | -756·75 (-7340·99 to 5827·50) |
| MHA detentions | 1030·18 (2806·08) | 586·34 (2922·25) | 513·38 (1673·99) | 280·07 (1068·47) | 1·29 (0·66 to 2·54) | 0·97 (0·39 to 2·43) | -19·38 (-732·64 to 693·88) |
| Total | 56943·54 (116602·67) | 32755·62 (79840·41) | 24993·61 (72900·16) | 13434·03 (46904) | 0·86 (0·51 to 1·44) | 0·80 (0·49 to 1·33) | -12614·60 (-35365·96 to 10136·75) |

CI=confidence interval; CMHT=Community Mental Health Teams; HTT=Home Treatment Team; IRR=incident rate ratio; MHA=Mental Health Act; SD=standard deviation
*p<0·05, **p<0·001
The IRRs presented are exponentiated coefficients of the interaction term – treatment*time – for each follow-up period· Estimated models at GLM with Gamma family and log link· Costs in £ at 2022/23 prices· Adjusted models included ethnicity, smoking status, and F6 diagnosis·

| Table S4: Other all-cause hospital use in pre- and post-index date periods presented as both medians and IQRs and means and SDs alongside results from unadjusted and adjusted negative binomial regression models | | | | | | |
|---|---|---|---|---|---|---|
| | Student (N=1 193) | | Control (N=3 508) | | | |
| | 6m pre | 6m post | 6m pre | 6m post | | |
| 6 months | *Median (IQR)* | *Median (IQR)* | *Median (IQR)* | *Median (IQR)* | | |
| Inpatient admissions | 0 (0 to 0) | 0 (0 to 0) | 0 (0 to 0) | 0 (0 to 0) | | |
| *Emergency admissions* | 0 (0 to 0) | 0 (0 to 0) | 0 (0 to 0) | 0 (0 to 0) | | |
| *Elective admissions* | 0 (0 to 0) | 0 (0 to 0) | 0 (0 to 0) | 0 (0 to 0) | | |
| Bed days | 0 (0 to 0) | 0 (0 to 0) | 0 (0 to 0) | 0 (0 to 0) | | |
| Emergency department attendances | 0 (0 to 1) | 0 (0 to 1) | 0 (0 to 1) | 0 (0 to 1) | | |
| Outpatient appointments | 0 (0 to 2) | 0 (0 to 2) | 0 (0 to 1·5) | 0 (0 to 2) | | |
| | *Mean (SD)* | *Mean (SD)* | *Mean (SD)* | *Mean (SD)* | *Unadjusted IRR (95% CI)* | *Adjusted IRR (95% CI)* |
| Inpatient admissions | 0·27 (1·94) | 0·26 (2·39) | 0·28 (1·59) | 0·30 (1·66) | 0·91 (0·67 to 1·23) | 0·90 (0·66 to 1·21) |
| *Emergency admissions* | 0·11 (0·41) | 0·09 (0·38) | 0·11 (0·45) | 0·12 (0·55) | 0·80 (0·54 to 1·17) | 0·78 (0·53 to 1·14) |
| *Elective admissions* | 0·16 (1·90) | 0·17 (2·29) | 0·13 (1·47) | 0·15 (1·52) | 0·96 (0·61 to 1·50) | 0·97 (0·62 to 1·51) |
| Bed days | 1·08 (5·64) | 0·76 (4·58) | 2·09 (12·45) | 2·66 (14·94) | 0·55 (0·36 to 0·84)* | 0·53 (0·35 to 0·81)* |
| Emergency department attendances | 0·84 (1·92) | 0·64 (1·64) | 0·69 (2·08) | 0·69 (3·25) | 0·75 (0·61 to 0·93)* | 0·74 (0·60 to 0·91)* |
| Outpatient appointments | 1·55 (3·79) | 1·61 (3·77) | 1·54 (4·35) | 1·61 (4·70) | 0·99 (0·80 to 1·22) | 0·99 (0·80 to 1·22) |
| | Student (N=1 072) | | Control (N=3 158) | | | |
| | 12m pre | 12m post | 12m pre | 12m post | | |
| 12 months | *Median (IQR)* | *Median (IQR)* | *Median (IQR)* | *Median (IQR)* | | |
| Inpatient admissions | 0 (0 to 0) | 0 (0 to 0) | 0 (0 to 0) | 0 (0 to 0) | | |
| *Emergency admissions* | 0 (0 to 0) | 0 (0 to 0) | 0 (0 to 0) | 0 (0 to 0) | | |
| *Elective admissions* | 0 (0 to 0) | 0 (0 to 0) | 0 (0 to 0) | 0 (0 to 0) | | |
| Bed days | 0 (0 to 1) | 0 (0 to 0) | 0 (0 to 1) | 0 (0 to 1) | | |
| Emergency department attendances | 1 (0 to 2) | 0 (0 to 1) | 0 (0 to 1) | 0 (0 to 1) | | |
| Outpatient appointments | 0 (0 to 3) | 1 (0 to 3·5) | 0 (0 to 3) | 0 (0 to 3) | | |
| | *Mean (SD)* | *Mean (SD)* | *Mean (SD)* | *Mean (SD)* | *Unadjusted IRR (95% CI)* | *Adjusted IRR (95% CI)* |
| Inpatient admissions | 0·52 (3·58) | 0·52 (4·89) | 0·60 (3·32) | 0·57 (3·23) | 1·06 (0·81 to 1·38) | 1·06 (0·81 to 1·38) |
| *Emergency admissions* | 0·19 (0·64) | 0·16 (0·64) | 0·24 (0·85) | 0·21 (0·81) | 0·93 (0·66 to 1·30) | 0·92 (0·66 to 1·28) |
| *Elective admissions* | 0·32 (3·51) | 0·35 (4·74) | 0·29 (3·13) | 0·31 (3·06) | 1·02 (0·70 to 1·49) | 1·02 (0·70 to 1·49) |
| Bed days | 2·09 (11·39) | 1·59 (9·25) | 4·23 (23·37) | 4·80 (26·46) | 0·67 (0·46 to 0·96)* | 0·66 (0·46 to 0·96)* |
| Emergency department attendances | 1·42 (2·58) | 1·19 (2·71) | 1·34 (4·08) | 1·26 (4·09) | 0·89 (0·74 to 1·08) | 0·90 (0·75 to 1·09) |
| Outpatient appointments | 2·88 (5·89) | 3·35 (7·38) | 3·03 (7·78) | 3·08 (7·46) | 1·14 (0·94 to 1·40) | 1·14 (0·94 to 1·40) |
| | Student (N=180) | | Control (N=529) | | | |
| | 5y pre | 5y post | 5y pre | 5y post | | |
| 5 years | *Median (IQR)* | *Median (IQR)* | *Median (IQR)* | *Median (IQR)* | | |
| Inpatient admissions | 0 (0 to 2) | 1 (0 to 2) | 1 (0 to 3) | 1 (0 to 3) | | |
| *Emergency admissions* | 0 (0 to 1) | 0 (0 to 1) | 0 (0 to 1) | 0 (0 to 1) | | |
| *Elective admissions* | 0 (0 to 1) | 0 (0 to 1) | 0 (0 to 1) | 0 (0 to 1) | | |
| Bed days | 1 (0 to 5) | 1 (0 to 4) | 2 (0 to 7) | 1 (0 to 8) | | |
| Emergency department attendances | 3 (1 to 5) | 2 (1 to 6) | 3 (1 to 6) | 2 (0 to 5) | | |
| Outpatient appointments | 5 (1·5 to 15) | 6·5 (2 to 21·5) | 5 (1 to 15) | 6 (1 to 17) | | |

| | *Mean (SD)* | *Mean (SD)* | *Mean (SD)* | *Mean (SD)* | *Unadjusted IRR (95% CI)* | *Adjusted IRR (95% CI)* |
|---|---|---|---|---|---|---|
| Inpatient admissions | 1·71 (3·22) | 1·79 (2·89) | 2·50 (5·56) | 2·82 (11·34) | 0·93 (0·61 to 1·42) | 0·96 (0·63 to 1·45) |
| *Emergency admissions* | 0·81 (1·81) | 0·67 (1·62) | 1·14 (2·80) | 1·04 (5·06) | 0·90 (0·53 to 1·52) | 0·96 (0·57 to 1·61) |
| *Elective admissions* | 0·85 (2·36) | 1·08 (2·25) | 1·09 (4·24) | 1·55 (9·64) | 0·89 (0·51 to 1·55) | 0·89 (0·51 to 1·55) |
| Bed days | 6·18 (13·55) | 6·67 (23·79) | 18·21 (99·32) | 20·44 (103·10) | 0·96 (0·54 to 1·71) | 0·99 (0·56 to 1·77) |
| Emergency department attendances | 4·53 (6·78) | 4·58 (6·29) | 6·34 (16·91) | 6·20 (24·79) | 1·03 (0·72 to 1·48) | 1·07 (0·75 to 1·52) |
| Outpatient appointments | 11·99 (18·66) | 16·95 (30·56) | 11·60 (19·03) | 13·85 (21·80) | 1·18 (0·83 to 1·69) | 1·19 (0·83 to 1·70) |

CI=confidence interval; IQR=interquartile range; IRR=incident rate ratio; SD=standard deviation
*p<0·05, **p<0·001
The IRRs presented are coefficients of the interaction term – treatment*time – for each follow-up period· Adjusted models included F6 diagnosis·

| Table S5· Impact of Recovery College enrolment on other all-cause hospital use costs | | | | | | | |
|---|---|---|---|---|---|---|---|
| | Student (N=1 193) | | Control (N=3 508) | | | | |
| | Mean (SD) | | Mean (SD) | | | | |
| Costs (£) | 6 months pre | 6 months post | 6 months pre | 6 months post | Unadjusted IRR (95% CI) | Adjusted IRR (95% CI) | Marginal Effects £ (95% CI) |
| Bed days | 431·31 (2247·45) | 323·4 (1883·82) | 875·13 (5168·25) | 1116·49 (6155·53) | 0·55 (0·32 to 0·93)* | 0·53 (0·31 to 0·91)* | -376·36 (-687·93 to -64·78)* |
| Emergency department attendances | 231·63 (525·74) | 176·17 (449·34) | 187·43 (566·64) | 188·51 (883·46) | 0·75 (0·58 to 0·99)* | 0·74 (0·56 to 0·98)* | -58·89 (-111·90 to -5·88)* |
| Outpatient appointments | 273·16 (665·03) | 282·24 (662·47) | 271·89 (764·7) | 283·49 (826·49) | 0·99 (0·78 to 1·25) | 0·99 (0·78 to 1·25) | -3·04 (-68·62 to 62·54) |
| Total | 936·1 (2726·05) | 781·81 (2268·17) | 1334·46 (5517·95) | 1588·5 (6614·2) | 0·68 (0·50 to 0·93)* | 0·67 (0·49 to 0·91)* | -443·04 (-792·03 to -94·04)* |
| | Student (N=1 072) | | Control (N=3 158) | | | | |
| | Mean (SD) | | Mean (SD) | | | | |
| Costs (£) | 12 months pre | 12 months post | 12 months pre | 12 months post | Unadjusted IRR (95% CI) | Adjusted IRR (95% CI) | Marginal Effects £ (95% CI) |
| Bed days | 905·58 (4604·51) | 654·12 (3670·28) | 1794·74 (9824·16) | 2063·14 (10850·8) | 0·67 (0·39 to 1·16) | 0·66 (0·38 to 1·15) | -435·91(-1044·06 to 172·24) |
| Emergency department attendances | 420·07 (926·47) | 327·55 (747·48) | 376·56 (1165·81) | 356·79 (1354·31) | 0·89 (0·71 to 1·13) | 0·90 (0·72 to 1·13) | -33·61 (-112·31 to 45·10) |
| Outpatient appointments | 518·03 (1041·16) | 563·32 (1247·17) | 542·39 (1350·92) | 537·06 (1293·06) | 1·15 (0·92 to 1·42) | 1·14 (0·92 to 1·42) | 74·27 (-46·41 to 194·95) |
| Total | 1843·68 (5226·28) | 1544·99 (4272·01) | 2713·69 (10550·6) | 2956·99 (11553·13) | 0·83 (0·61 to 1·13) | 0·82 (0·59 to 1·12) | -412·43 (-1084·95 to 260·09) |
| | Student (N=180) | | Control (N=529) | | | | |
| | Mean (SD) | | Mean (SD) | | | | |
| Costs (£) | 5 years pre | 5 years post | 5 years pre | 5 years post | Unadjusted IRR (95% CI) | Adjusted IRR (95% CI) | Marginal Effects £ (95% CI) |
| Bed days | 5129·73 (25631·66) | 3448·46 (19543·12) | 8163·51 (35179·73) | 6248·6 (31556·31) | 0·96 (0·40 to 2·32) | 0·99 (0·42 to 2·35) | -465·50 (-5465·83 to 4534·83) |
| Emergency department attendances | 1620·95 (2999·59) | 1055·86 (3129·57) | 1701·71 (4441·47) | 1106·83 (4095·63) | 1·03 (0·62 to 1·71) | 1·08 (0·63 to 1·85) | 101·46 (-750·05 to 952·97) |
| Outpatient appointments | 2141·9 (3358·26) | 1799·86 (3480·09) | 2321·21 (4674·46) | 1707·92 (3373·6) | 1·18 (0·80 to 1·76) | 1·19 (0·79 to 1·78) | 480·37 (-545·97 to 1506·71) |
| Total | 8892·57 (27447·88) | 6304·18 (21609·11) | 12186·43 (37309·43) | 9063·35 (33685·73) | 1·07 (0·61 to 1·86) | 1·10 (0·64 to 1·90) | 112·80 (-5364·83 to 5590·43) |

CI=confidence interval; IRR=incident rate ratio; SD=standard deviation
*p<0·05, **p<0·001
The IRRs presented are exponentiated coefficients of the interaction term – treatment*time – for each follow-up period· Estimated models at GLM with Gamma family and log link· Costs in £ at 2023/24 prices· Adjusted models included F6 diagnosis·

**Table S6· Impact of Recovery College enrolment on HoNOS scores: Before-and-after study in students only**

| | 6 months (N=475) | | | 12 months (N=793) | | | 5 years (N=979) | | |
|---|---|---|---|---|---|---|---|---|---|
| | *Pre-index date* | *Post--index date* | *Change over time* | *Pre-index date* | *Post--index date* | *Change over time* | *Pre-index date* | *Post--index date* | *Change over time* |
| *HoNOS item* | *Median (IQR)* | *Median (IQR)* | *IRR or OR (95% CI)* | *Median (IQR)* | *Median (IQR)* | *OR (95% CI)* | *Median (IQR)* | *Median (IQR)* | *OR (95% CI)* |
| 1. Agitated behaviour | 0 (0 to 1) | 0 (0 to 1) | *OR=0·70 (0·55 to 0·91) | 0 (0 to 1) | 0 (0 to 1) | *OR=0·75 (0·62 to 0·91) | 0 (0 to 1) | 0 (0 to 1) | *OR=0·76 (0·63 to 0·91) |
| 2. Self-injury | 0 (0 to 1) | 0 (0 to 0) | OR=0·82 (0·61 to 1·09) | 0 (0 to 1) | 0 (0 to 0) | *OR=0·76 (0·61 to 0·96) | 0 (0 to 1) | 0 (0 to 0) | **OR=0·63 (0·51 to 0·78) |
| 3. Problem drinking or drugs | 0 (0 to 1) | 0 (0 to 1) | OR=1·02 (0·77 to 1·36) | 0 (0 to 1) | 0 (0 to 1) | OR=0·96 (0·77 to 1·20) | 0 (0 to 1) | 0 (0 to 1) | OR=0·93 (0·77 to 1·14) |
| 4. Cognitive problems | 0 (0 to 1) | 0 (0 to 1) | OR=0·91 (0·69 to 1·20) | 0 (0 to 1) | 0 (0 to 1) | OR=0·97 (0·79 to 1·19) | 0 (0 to 1) | 0 (0 to 1) | OR=0·96 (0·80 to 1·15) |
| 5. Physical illness | 0 (0 to 1) | 0 (0 to 1) | OR=1·10 (0·86 to 1·40) | 0 (0 to 1) | 0 (0 to 2) | OR=1·19 (0·99 to 1·44) | 0 (0 to 1) | 0 (0 to 2) | *OR=1·25 (1·06 to 1·48) |
| 6. Hallucinations | 0 (0 to 2) | 0 (0 to 1) | *OR=0·75 (0·59 to 0·97) | 0 (0 to 2) | 0 (0 to 1) | *OR=0·81 (0·66 to 0·98) | 0 (0 to 2) | 0 (0 to 1) | *OR=0·78 (0·66 to 0·93) |
| 7. Depressed mood | 1·5 (1 to 2) | 1 (1 to 2) | *OR=0·74 (0·59 to 0·93) | 1 (1 to 2) | 1 (1 to 2) | OR=0·87 (0·73 to 1·04) | 1 (1 to 2) | 1 (1 to 2) | *OR=0·83 (0·71 to 0·98) |
| 8. Other mental problems | 2 (1 to 3) | 2 (1 to 2) | *OR=0·76 (0·60 to 0·95) | 2 (1 to 3) | 2 (1 to 2) | *OR=0·77 (0·65 to 0·92) | 2 (1 to 3) | 2 (1 to 3) | *OR=0·84 (0·72 to 0·99) |
| 9. Relationship problems | 1 (0 to 2) | 1 (0 to 2) | OR=0·93 (0·74 to 1·16) | 1 (0 to 2) | 1 (0 to 2) | OR=0·91 (0·77 to 1·09) | 1 (0 to 2) | 1 (0 to 2) | OR=0·89 (0·76 to 1·04) |
| 10. Daily living problems | 1 (0 to 2) | 0 (0 to 1) | OR=0·86 (0·67 to 1·09) | 1 (0 to 2) | 0 (0 to 1) | OR=0·89 (0·74 to 1·07) | 1 (0 to 1) | 0 (0 to 1) | OR=0·91 (0·77 to 1·08) |
| 11. Living conditions | 0 (0 to 1) | 0 (0 to 1) | OR=0·90 (0·69 to 1·16) | 0 (0 to 1) | 0 (0 to 1) | OR=0·93 (0·76 to 1·14) | 0 (0 to 1) | 0 (0 to 1) | OR=0·96 (0·80 to 1·16) |
| 12. Occupational problems | 1 (0 to 2) | 1 (0 to 2) | OR=0·81 (0·64 to 1·02) | 1 (0 to 2) | 1 (0 to 2) | *OR=0·79 (0·66 to 0·95) | 1 (0 to 2) | 1 (0 to 2) | OR=0·88 (0·75 to 1·04) |

HoNOS=Health of the Nation Outcome Scale; IQR=interquartile range; IRR=incident rate ratio; OR=odds ratio
*p<0·05, **p<0·001
The overall change in HoNOS scores was evaluated using negative binomial regression, accounting for data overdispersion· Individual HoNOS items were examined through ordinal regression to appropriately reflect their ordinal scale·

| Table S7: Impact of Recovery College enrolment on mental health service use: Before-and-after study in student group | | | |
|---|---|---|---|
| | 6 months<br>Student N=1 435 | 12 months<br>Student N=1 293 | 5 years<br>Student N=206 |
| | *IRR (95% CI)* | *IRR (95% CI)* | *IRR (95% CI)* |
| Inpatient admissions | 0·55 (0·45 to 0·68)** | 0·58 (0·47 to 0·72)** | 0·69 (0·42 to 1·14) |
| Bed days | 0·66 (0·44 to 1·00) | 0·72 (0·48 to 1·07) | 0·59 (0·25 to 1·35) |
| HTT events | 0·50 (0·34 to 0·74)* | 0·58 (0·40 to 0·83)* | 0·67 (0·33 to 1·37) |
| Liaison psychiatry | 0·49 (0·37 to 0·65)** | 0·60 (0·47 to 0·77)** | 0·89 (0·55 to 1·44) |
| CMHT events | 0·99 (0·89 to 1·09) | 1·02 (0·92 to 1·14) | 0·82 (0·63 to 1·06) |
| MHA detentions | 0·61 (0·48 to 0·79)** | 0·73 (0·57 to 0·94)* | 0·80 (0·42 to 1·50) |

CI=confidence interval; CMHT=Community Mental Health Team; HTT= Home Treatment Team; IRR=incident rate ratio; MHA=Mental Health Act
*p<0·05, **p<0·001

| Table S8: Impact of Recovery College enrolment on other all-cause hospital use: Before-and-after study in student group | | | |
|---|---|---|---|
| | 6 months<br>Student N=1 193 | 12 months<br>Student N=1 072 | 5 years<br>Student N=180 |
| | *IRR (95% CI)* | *IRR (95% CI)* | *IRR (95% CI)* |
| Inpatient admissions | 0·97 (0·75 to 1·26) | 1·00 (0·79 to 1·27) | 1·05 (0·76 to 1·46) |
| Emergency admissions | 0·84 (0·61 to 1·15) | 0·81 (0·61 to 1·08) | 1·08 (0·70 to 1·67) |
| Elective admissions | 1·08 (0·74 to 1·58) | 1·11 (0·80 to 1·54) | 0·82 (0·53 to 1·28) |
| Bed days | 0·70 (0·50 to 0·98)* | 0·76 (0·56 to 1·02) | 1·27 (0·84 to 1·91) |
| Emergency department attendances | 0·76 (0·64 to 0·90)* | 0·83 (0·72 to 0·97)* | 1·23 (0·87 to 1·75) |
| Outpatient appointments | 1·03 (0·87 to 1·22) | 1·16 (0·98 to 1·37) | 1·41 (1·05 to 1·90)* |
| CI=confidence interval; IRR=incident rate ratio<br>*p<0·05, **p<0·001 | | | |
